# Racial/Ethnic Inequalities for Legal Intervention Injuries Treated in U.S. Emergency Departments: United States (2004-2021)

**DOI:** 10.1101/2025.05.09.25327323

**Authors:** Mina Kim, Justin M. Feldman, Phillip Atiba Solomon

**Author notes:** CORRESPONDING AUTHOR Mina Kim, Center for Policing Equity, 8605 Santa Monica Blvd, PMB 54596, West Hollywood, CA 90069-4109.

## Abstract

Nonfatal injuries caused by law enforcement are a widespread yet underexamined public health concern in the United States, with significant implications for racial health equity. Using data from the National Electronic Injury Surveillance System—All Injury Program (NEISS-AIP) from 2004 to 2021, this study estimated national trends in emergency department visits for injuries inflicted by on-duty police and assessed disparities across racial and ethnic groups. The analysis incorporated imputation for missing data on race and parametric bootstrapping to account for sampling and imputation uncertainty. We found that legal intervention injury rates remained relatively stable over time, with Black individuals consistently experiencing rates over five times higher than White individuals, and Latinx individuals showing moderately higher rates than Whites with some evidence of decline. These findings suggest that, despite heightened public scrutiny and advocacy following nationwide protests against racialized policing, rates of nonfatal injuries from law enforcement have remained high and unequally distributed by race.

## Introduction

In the United States, injuries caused by law enforcement are a public health concern and driver of racial health inequities.^1–4^ While fatalities have attracted considerable public attention, nonfatal injuries inflicted by police are far more prevalent and therefore more typical in terms of adverse police-related outcomes. Analyzing non-fatal injury trends can therefore help to answer critical questions about how populations experience policing, including whether the major advocacy efforts related to racial inequity in policing occurring from 2014-2021^5^ coincided with changes to rates of police-related nonfatal injury overall or by racial and ethnic group.

## Methods

This repeated cross-sectional study analyzed data from the National Electronic Injury Surveillance System—All Injuries Program (NEISS-AIP), a nationally representative sample of US hospital emergency departments (EDs), for the period 2004-2021. For NEISS, coders assign each injury an intent category; “legal intervention” includes an injury/poisoning caused by on-duty police or other legal authorities, including private security guards. NEISS-AIP offers an advantage over administrative claims data, which underreport substantial shares of legal intervention injuries.^6^ We used NEISS’s pre-defined race and ethnicity categories, which are derived from medical records: African American/Black (hereafter, Black), including Black Hispanic/Latinx individuals; White (excluding Hispanic White); Hispanic/Latinx (non-Black persons of any race); and other races (non-Hispanic persons who were American Indian/Alaska Native, Asian American, Pacific Islander, or multiracial). We fit quasi-Poisson models for injury rates, treating year as a spline (for visualization) or as linear (to assess trends quantitatively). To construct 95% confidence intervals, we used parametric bootstrapping, propagating error from imputation (used for the 22% of patients with missing race) and survey design. Additional methodological details are available in the eAppendix in Supplement 1.

## Results

Between 2004 and 2021, a total of 1,500,577 ED visits (95% CI: 1073632, 1927522) in the US were for legal intervention injuries (patient sex: 85% men (95% CI: 60, 100); age: mean 33 [SD 12]. For patients with non-missing race/ethnicity data, 41% were White (95% CI: 31, 51), 42% were Black (95% CI: 21, 63), and 14% were Latinx (95% CI: 5, 22). Most injuries were relatively minor, with 4% (95% CI: 3, 6) requiring hospitalization.

Legal intervention injury rates for the US population as a whole remained relatively stable over the study period (Figure 1), with the 2021 rate at 92% (95% CI: 71, 119) the level of the 2004 rate. Averaged over the study period, injury rates for Black people were 5.30 times that of White people (95 % CI: 4.55, 6.15). Rates for Latinx people were 1.45 times that of White people (95 % CI: 1.21, 1.72). Point estimates suggested a decrease in the Black-White rate ratio (RR) of 6%— from 5.48 in 2004 to 5.13 in 2021— but uncertainty was high, with a 95% confidence interval ranging from a 42% reduction to a 51% increase. For the Latinx population, evidence suggests the RR decreased by 45% (95% CI: 12%, 66%).

**Figure 1:**
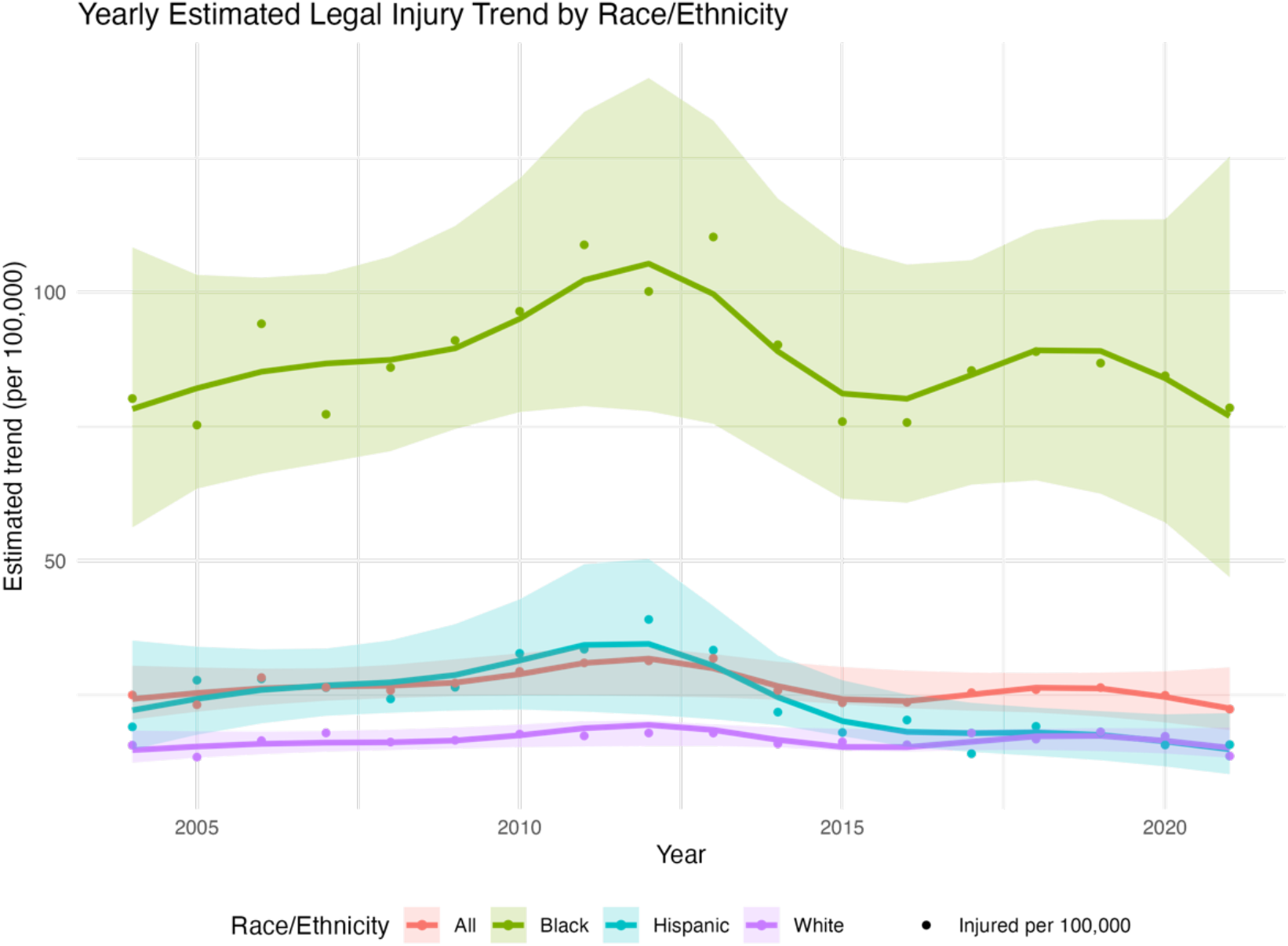
Yearly estimated legal intervention injury trend by race/ethnicity

## Discussion

Despite expectations that policy responses to major protests against racialized police violence might have led to substantial change, our findings suggest any shifts in the rate of nonfatal legal intervention injuries for the overall population were relatively modest, ranging from a decrease by approximately one-third to an increase of one-fifth. Our findings of a decreasing Latinx-White RR should be interpreted cautiously, as misclassification of Latinx patients in US medical data is high and may vary as EDs with different classification practices enter and exit the NEISS-AIP sample. While we cannot draw conclusions about the direction of change to the Black-White RR, it is clear that it remained persistently high over the entire study period, with point estimates always exceeding 5.

**Table 1:** Legal intervention injuries in hospital emergency departments, 2004-2021.

|  |  | Total Number of ED visits<br>[95% CI] | % of legal intervention injuries<sup>1</sup><br>[95% CI] |
| --- | --- | --- | --- |
| *Sex*<br>(Unknown/Non-binary=0.02%)<sup>2</sup> |  |  |  |
|  | Men | 1270350 (896648, 1644051) | 84.68% [59.77%, 100.00%]<sup>3</sup> |
|  | Women | 229863 [173663, 286062] | 15.32% [11.58%, 19.07%] |
| *Race/ethnicity<sup>4</sup>*<br>(Unknown=22.08%) |  |  |  |
|  | White | 480110 [358368, 601852] | 41.06% [30.65%, 51.47%] |
|  | Black | 494432 [247922, 740942] | 42.29% [21.20%, 63.37%] |
|  | Latinx<br>(non-Black) | 161917 [62633, 261201] | 13.85% [5.36%, 22.34%] |
|  | Other races | 32777 [12232, 53321] | 2.80% [1.05%, 4.56%] |
| *Age group*<br>(Unknown=0.06%) |  |  |  |
|  | 14 and under | 26349 [15453, 37246] | 1.76% [1.03%, 2.48%] |
|  | 15 to 24 | 424842 [302945, 546739] | 28.33% [20.20%, 36.46%] |
|  | 25 to 34 | 469617 [334934, 604300] | 31.31% [22.33%, 40.30%] |
|  | 35 to 44 | 312805 [216562, 409048] | 20.86% [14.44%, 27.28%] |
|  | 45 to 54 | 182773 [130607, 234939] | 12.19% [8.71%, 15.67%] |
|  | 55 and above | 83274 [61688, 104859] | 5.55% [4.11%, 6.99%] |
| *Disposition of case*<br>(Unknown=0.01%) |  |  |  |
|  | Treated and released | 1335886 [972719, 1699054] | 89.03% [64.83%, 113.23%] |
|  | Transferred and released | 27617 [19403, 35830] | 1.84% [1.29%, 2.39%] |
|  | Hospitalized | 64505 [38009, 91002] | 4.30% [2.53%, 6.06%] |
|  | Observation | 46236 [-34468, 126939] | 3.08% [0.00%, 8.46%]<sup>3</sup> |
|  | Left against medical advice/<br>Without being seen | 26245 [11195, 41294] | 1.75% [0.75%, 2.75%] |
| *Immediate cause of injury*<br>(Unknown=0.90%) |  |  |  |
|  | Cut/pierce | 141651 [41847, 241455] | 9.53% [2.81%, 16.24%] |
|  | Dog bite | 70931 [33307, 108555] | 4.77% [2.24%, 7.30%] |
|  | Fall | 214262 [152156, 276368] | 14.41% [10.23%, 18.59%] |
|  | Firearm gunshot | 19285 [3701, 34869] | 1.30% [0.25%, 2.34%] |
|  | Struck by/against | 817528 [591204, 1043851] | 54.98% [39.76%, 70.20%] |
|  | Other | 223345 [167256, 279434] | 15.02% [11.25%, 18.79%] |
| Total |  | 1500577 [1073632, 1927522] |  |

<sup>1</sup> The denominator only includes legal intervention injuries with non-missing data for the respective variable<sup>2</sup> Non-binary category was only available in the 2021 data<sup>3</sup> 95% Confidence intervals for percentage that extend above 100% or below 0% have been truncated.<sup>4</sup> While “White” excludes Hispanic/Latinx, “Black” includes Latinx individuals. “Other races” include Asian, Pacific Islander, American Indian/Alaska Native, and more than one race.

## Data Availability

All data produced in the present study are available upon reasonable request to the authors

https://github.com/mak791/NEISS

## Notes

### Competing Interest Statement

The authors have declared no competing interest.

### Funding Statement

This study did not receive any funding

### Author Declarations

have followed all appropriate research reporting guidelines, such as any relevant EQUATOR Network research reporting checklist(s) and other pertinent material, if applicable.

