## Supplement 1 for "Racial/Ethnic Inequalities for Legal Intervention Injuries Treated in U.S. Emergency Departments: United States (2004-2021)"

### NEISS-AIP Data

We obtained open-access data from the National Electronic Injury Surveillance System - All Injury Program (NEISS-AIP) from the ICPSR repository (<https://www.icpsr.umich.edu/web/ICPSR/series/198>) for the period 2004-2021. NEISS-AIP is a stratified, random sample of US hospital emergency departments (EDs). EDs enter and exit the sample over time, and the number of EDs included in the sample varies as well. The mean number of EDs sampled annually during the study period was 51.

Identifiers referring to unique EDs were not available in the open-access NEISS-AIP dataset; instead, the data included primary sampling unit (PSU) identifiers. PSU identifiers are static, remaining the same even when one ED is replaced with another in the PSU.

To create unique ED identifiers, we used a table (available from the Consumer Product Safety Commission upon request, and included in the GitHub repository for this manuscript) that lists PSU IDs and the dates when the ED was replaced for each PSU.

### US Population Data

We obtained annual data for the US residential population (50 states and the District of Columbia) for 2004-2021, stratified by race/ethnicity, from the US Census Population Estimates Program (<https://www.census.gov/programs-surveys/popest/data/data-sets.html>). These are postcensal estimates that Bureau staff updates annually using data on births, deaths, and migration.

### Analysis Software

We conducted all analyses in R version 4.4. All models were generalized additive models (GAMs) fit with the mgcv package using the gam() function.

### Race Imputation

We imputed race/ethnicity for the 22% of injuries with missing race and/or ethnicity data using the following steps:

1. Fit a multinomial GAM to the injuries with non-missing patient race/ethnicity. The response variable was race category: African American/Black (Hispanic and non-Hispanic), non-Hispanic White, Hispanic/Latinx excluding Black, and other non-Hispanic races. The model included patient-level predictors: age, sex; hospital stratum; ED random intercepts; and ED-level summary measures of all injuries reported to NEISS-AIP by the treatment ED (regardless of intent and pooled across all available years of data for that ED): mean monthly count of injuries, % of injuries classified as assault, % of injuries classified as legal intervention, mean patient age, proportion of injuries for which the treated patient was male. All continuous variables were fit as penalized splines.
2. Use the multinomial GAM fit to generate race/ethnicity predictions for each patient with missing race/ethnicity. This yielded four predicted probabilities for each patient (i.e., the probability that the patient belonged to each of the 4 racial/ethnic groups).
3. Create values across 4 race/ethnicity variables for each patient regardless of missingness. For patients with non-missing race, assign 1 for their reported race, and 0 for all other races. For patients with missing race, assign their model-predicted probabilities for each race/ethnicity category.
4. Using the survey package to account for survey weights and design (with the NEISS-AIP’s PSU and stratum variables specified), calculate aggregate injury counts as a weighted sum of each of the 4 race categories by year, and estimate the variance for each of these counts.

### Outcome Models

We fit four GAMs to model the annual, population-based rate of legal intervention injuries: one pair was fit for the overall population and the other pair was race-stratified. Within each pair, one model included a linear time trend (year treated as a continuous variable), and the other treated year as a penalized spline. All four GAMs were fit with a quasi-Poisson family and log link, had a response variable equal to the annual count of legal intervention injuries, and included a ln(population) offset referring to that year’s US population (total or race-specific).

The race-stratified models additionally included an indicator variable for race and a race*year interaction term. In the model that treated time as nonlinear, this was specified as a factor-smooth interaction.

All reported analytic results were based on rates predicted from these models. For example, to calculate the change in Black-white rate ratio we used the race-stratified model with a linear time trend to predict injury rates for each race*year. Then, we calculated the ratio of Black-to-white rates based on their model predictions in 2004. We compared this to the rate ratio for 2021, also calculated based on the model-predicted rates. Confidence intervals for these results were based on the procedure described below.

### Estimation of Uncertainty

NEISS-AIP’s survey design variables only allow for an analytic approach to variance estimation for injury *counts* rather than for population-based injury *rates*. We therefore developed a multi-stage procedure to generate the 95% confidence intervals for all analytic results that allowed us to propagate uncertainty from both survey design and imputation in our outcome models.

The procedure included a combination of parametric bootstrapping and the delta method. For the race-specific estimates, we took the following steps:

1. Start with the annual, race-specific injury counts and variance estimates calculated using the method described in the Race Imputation section above. Note that these variance estimates reflect uncertainty due to sampling and survey design only (var(survey)).
2. Use parametric bootstrapping (n = 10,000 iterations) to estimate the additional variance attributable to the imputation process. In each iteration, every injury case with missing patient race is randomly assigned a single race by drawing from a multinomial distribution, which is parameterized by the model-predicted, patient-specific probabilities for each racial category (e.g., a patient with a predicted probability of 0.5 for Black would be expected to be assigned Black in half of bootstrapped replicates). Race for those with non-missing data remains unchanged.
3. Calculate weighted injury counts by race and year for each of the bootstrap iterations as the survey-weighted sum of each racial category. Every race*year combination will have a set of 10,000 replicate count values, and the variance of each set is the variance attributable to imputation (var(imputation)) for the race*year combination.
4. Add var(survey) to var(imputation) for each race*year combination. We assume independence, and the delta method formula is therefore a simple sum: var(total) = var(survey) + var(imputation).
5. Perform an additional parametric bootstrap (n = 10,000) to generate a set of race-year injury counts whose distributions are parameterized to reflect each point estimate along with the newly calculated total variance. This is done by drawing a random number from a gamma distribution (shape = point_estimate^2/var_total; rate = point_estimate/var_total) for each race*year combination.
6. Fit each model to each of the bootstrap replicates produced in Step 5. The population offset is treated as a fixed value and does not change across replicates. Calculate 95% confidence limits as the 2.5th and 97.5th percentiles of model results.

The procedure for the overall (not race-stratified) counts was similar, but did not need to account for error from imputation. Instead, we simply generated weighted annual injury counts and variance estimates with the survey package, then used parametric bootstrapping based on a gamma distribution parameterized as described in Step 5.
